# Multiplex ACE2-RBD Binding Inhibition Assay: An Integrated Tool for Assessing Neutralizing Antibodies to SARS-CoV-2 Variants and Protection against Breakthrough Infections

**DOI:** 10.1101/2024.10.25.24316126

**Authors:** Emma Bloch, Laura Garcia, Françoise Donnadieu, Jason Rosado, Delphine Planas, Timothée Bruel, Laurent Hocqueloux, Thierry Prazuck, Olivier Schwartz, Laura Tondeur, Laurie Pinaud, Arnaud Fontanet, Stéphane Pelleau, Michael White

## Abstract

SARS-CoV-2 remains a significant health threat due to its high infection and mutation rates. The emergence of new variants of concern poses challenges as they can lead to immune escape mutations, potentially reducing the efficacy of vaccines and antibody therapeutics. The receptor binding domain (RBD) of SARS-CoV-2 is particularly noteworthy as it is both the most rapidly evolving domain and the principal target of neutralizing antibodies. As an alternative to time-consuming and expensive neutralization assays, we have developed a bead-based multiplex surrogate virus neutralization test based on ACE2-RBD binding inhibition. We demonstrated how our high-throughput assay allows us to simultaneously assess anti-RBD neutralizing antibodies levels against multiple SARS-CoV-2 variants, providing data that is consistent with the gold-standard live virus neutralization assay. The utility of this assay was demonstrated by applying it to a large French population cohort to demonstrate that hybrid immunity (generated by a combination of vaccination and infection) is associated with protection against infection with the Omicron BA.1 and BA.2 lineages of SARS-COV-2.

## Introduction

SARS-CoV-2 is an enveloped RNA virus with surface projections that give rise to its corona appearance. Its genome encodes four structural proteins: spike (S), nucleocapsid (N), membrane (M), and envelope (E)^1^. The S and N proteins are highly immunogenic, eliciting a rapid immune response. The S glycoprotein is composed of two subunits, which form a trimeric structure on the virus surface. Subunit one (S1) harbors the receptor binding domain (RBD) of the virus, which binds to the angiotensin-converting enzyme-2 (ACE2) receptor on the host cell surface. Subunit two (S2) enables the fusion of the viral envelope with the host cellular membrane.

SARS-CoV-2 enters the organism mainly through the nasopharyngeal and oropharyngeal tracts of the host, via the cellular angiotensin-converting-enzyme-2 (ACE2) receptor. Cell entry also requires the presence of the transmembrane protease serine 2 (TMPRSS2) which cleaves the S protein and thus enables the membrane fusion and internalization^2^. The receptor involved in the cell entry plays a crucial role in determining the viral tropism and influences the severity of infection^3^. Primary infection or vaccine immunization leads to immune memory, providing protective immunity against subsequent infections. Various types of immunoglobulins targeting SARS-CoV-2 proteins are produced at different stages post-infection^4^. However, the concentration of these immunoglobulins varies significantly among individuals and over time, influenced by factors such as isotype, targeted antigen, and previous infection or vaccination history^5^.

Neutralizing antibodies interfere with the cell entry mechanism primarily by blocking the interaction of the RBD with the human cell receptor ACE2^6^. Anti-RBD antibodies account for approximately 90% of the neutralizing activity^7^. Several studies have demonstrated that the neutralizing antibody response to SARS-CoV-2 provides a correlate of protection^8,9^. Therefore, understanding the longevity and kinetics of the neutralizing antibody response is crucial as it has significant implications for immune protection and for the development of effective vaccination strategies.

Since the virus’s emergence, thousands of cumulative mutations have occurred, primarily within the S protein followed by the N protein, while the M and E proteins show a lower mutation rate in the SARS-CoV-2 genome^10^. These mutations can lead to evolutionary advantages, such as immune escape, higher transmission rate, varying severity of infection, and impact on the performance of diagnostic tools (i.e., antigenic tests), vaccines and therapeutic medicines. The World Health Organization (WHO) has been monitoring SARS-CoV-2 evolution since January 2020, categorizing variants into variants of interest (VOIs) and variants of concern (VOCs). VOCs, including Alpha (B.1.1.7.), Beta (B.1.351), Gamma (P.1), Delta (P.1.617.2), and Omicron (multiple lineages), are linked to increased transmissibility, virulence, or reduced effectiveness of public health measures. The study and monitoring of emerging virus strains is especially important for vaccine development strategies, since first-generation vaccines elicited an immune response against the original Wuhan-Hu-1 isolate (wild-type).

The gold standard method to measure the neutralization capacity of antibodies remains the live virus neutralization assay, a method based on living viruses and cell culture, which is time-consuming, does not allow the multiplexing of several variants, and requires a level 3 biosafety laboratory. Additionally, highly trained operators and biosafety containment are required. Several alternative methods have been explored, including pseudo-based virus neutralization assays, ELISA-based assays, and surrogate virus neutralization assays. To better understand the neutralizing antibody response to SARS-CoV-2 in large population studies, we developed a multiplex surrogate virus neutralization assay. This assay mimics the virus-host interaction in vitro, based on the interaction between the SARS-CoV-2 RBD and ACE2^11,12^. The assay will assess the functional inhibition of the ACE2-RBD binding, enabling us to predict protection to a breakthrough infection and vaccine effectiveness at population level.

## Material and Methods

### Patient cohort and Sample collection

#### Longitudinal cohort - COVID-Oise study

From May 2020 to May 2022, 900 individuals from the town of Crépy-en-Valois in the Oise Department in France were surveyed. In winter 2020, scientists at Institut Pasteur initiated a longitudinal cohort study, named the COVID-Oise cohort ^13,14^. Participants comprised a wide age range (5–101 years), ranging from children to nursing home residents. The inclusion criteria were to live, work and/or study in the town of Crépy-en-Valois (ca. 15,000 inhabitants) at the time of study initiation. No exclusion criteria were applied. Participants were invited four times for collection of epidemiological data and serum samples. Data and samples from sessions held in November 2020 (Session 1), April 2021 (Session 2), November 2021 (Session 3) and May 2022 (Session 4) were used in this analysis. The cohort included men and women of all ages both uninfected and infected with SARS-CoV-2, as well as some vaccinated individuals (1 doses up to 4 doses).

#### Convalescent and vaccinated individuals - Orléans cohort

A longitudinal clinical study was conducted in Orléans (France), enrolling 170 individuals who had PCR-confirmed SARS-CoV-2 infection with varying levels of disease severity. Additionally, blood samples were collected from 30 healthy individuals as negative controls. The study aimed to describe the persistence of specific and neutralizing antibodies over a 24-month period starting from August 2020. At enrollment, written informed consent was collected and participants completed a questionnaire covering sociodemographic characteristics, virological findings (SARS-CoV-2 RT–qPCR results), clinical data (date of symptom onset, type of symptoms, hospitalization) and data related to anti-SARS-CoV-2 vaccination if applicable (vaccine brand, date of each vaccination dose). Serological status of participants was assessed every 3 months^15^.

A subset of individuals who underwent anti-SARS-CoV-2 vaccination had weekly blood sampling after the first dose of vaccine for a period of 52 weeks post vaccination. Samples were collected weekly for the first 6 weeks, then once every two weeks from week 6 to 24, then every month until week 52. These 12 individuals received a first dose of vaccine at week 0, a second dose at week 4 and a third booster dose at between week 24 and 32. These samples include men and women of all ages, some of whom got infected with SARS-CoV-2 Delta or Omicron variant (lineages BA.1 or BA.2). A total of 253 sera samples were collected.

### Antigens – Recombinant proteins

A panel of 13 SARS-CoV-2 antigens have been used for the development of these multiplex surrogate neutralization tests, including Spike and RBD proteins from the ancestral virus (WT) and variants of concern (Alpha, Beta, Gamma, Delta and Omicron). Additionally, the RBD protein from the variant of interest, Kappa, has been included. Furthermore, during the preliminary phase of development, tests were conducted using SARS-CoV-2 N and M-E fusion proteins, as well as the spike (S) proteins from seasonal coronaviruses, including HCoV-NL63, HCoV-OC43, HCoV-229E, and HCoV-HKU1. The proteins used were either purchased from a Native Antigen (Oxford, UK) or produced at Institut Pasteur (Paris, France).

### Coupling antigens on beads

We used 1.25×10^6^ Luminex® magnetic beads to prepare 500µL of antigen coupled beads. Beads were vortexed and sonicated prior to being transferred to a 1.5mL microcentrifuge tube. A magnetic rack was used to remove the supernatant before washing the beads with Milli-Q water. Then, the beads were activated using 0.1M sodium phosphate (NaP) pH 6.2, 10 mg/mL of EDC (1-ethyl-3-[3-dimethylaminutesopropyl] carbodiimide hydrochloride) and 10 mg/mL sulfo-NHS (sulfo N-hydroxylsulfosuccinimide), they were incubated on a rotor in the dark for 20 min, at room temperature. This activation step allows the coupling of antigens to magnetic beads, through a covalent bond formed between a stable ester on the surface of the bead and the primary amine of the antigen. Thereafter, a magnetic rack was used to remove the supernatant before the beads were washed twice with PBS 1X (Phosphate-buffered saline). The mass of proteins coupled onto the beads was optimized previously, it was tested using a pool of 27 serum from RT-qPCR-confirmed SARS-CoV-2 patients and validated by generating a log-linear standard curve. Antigens were coupled to beads to their optimum concentration. The beads and antigens were incubated in a PBS 1X buffer on a rotor in the dark for 2 hours, at room temperature. Finally, the antigen coupled beads were washed three times with PBS-TBN (PBS, 1% BSA, 0.02% sodium azide and 0.05% Tween-20) and stored (using the same PBS-TBN buffer) at 4 ºC.

### Serological assay

A previously described 30-plex bead-based assay was used for simultaneous detection of IgG antibodies to 16 SARS-CoV-2 antigens (i.e., S proteins, RBD proteins, N protein, ME fusion protein), seasonal coronaviruses (i.e., Spike and NP proteins of HCoV-NL63, HCoV-OC43, HCoV-229E and HCoV-HKU1) and other antigens related to vaccine-preventable disease (i.e., Measles, Mumps, rubella)^16^. For this study purpose, we used the data generated previously for anti-NP, anti-Spike WT, anti-RBD for all variants (WT, Alpha, Beta, Gamma, Delta and Omicron) IgG antibodies. The protocol used was previously described^17^. Plates were read using a Luminex® MAGPIX® system and the median fluorescence intensity (MFI) was measured. On each assay plate, a blank (only with beads no serum) was included to control for background signal, as well as a standard curve prepared from two-fold serial dilutions (1/50 to 1/102,400) of a pool of 27 serum from RT-qPCR-confirmed SARS-CoV-2 patients. A 5-parameter logistic curve was used to convert MFI to relative antibody unit (RAU), relative to the standard curve performed on the same plate to account for inter-assay variations.

### Surrogate neutralization test based on the ACE2-RBD binding inhibition

To better understand the neutralizing antibody responses to SARS-CoV-2, we developed a multiplex surrogate virus neutralization test based on the functional inhibition of ACE2-RBD binding. In preliminary tests, the optimum ACE2, serum, and Streptavidin-Phycoerythrin concentration were selected. Subsequently, we have evaluated which SARS-CoV-2 antigens are capable of binding to ACE2. Antigens such as the M-E protein, S2 protein or N protein were excluded by their inability to bind to ACE2. Additionally, we study the possible cross-reactivity between various seasonal coronavirus S proteins (i.e., HCoV-NL63, HCoV-OC43, HCoV-229E and HCoV-HKU1). After analysis, we observed the ACE2 protein is unable to recognize and bind to seasonal coronavirus S protein. Lastly, the potential impact of multiplexing was assessed by testing each antigen-coupled bead individually and in a multiplex setting. No discernible interactions were observed among the different antigen-coupled beads during multiplexing. This trial enabled a transition from monoplex to multiplex.

Following assay optimization, an 8-plex bead-based assay including Spike and RBD proteins from the ancestral virus (WT) and RBD protein from variants of concern: Alpha, Beta, Gamma, Delta and Omicron (lineage BA.1) and variant of interest: Kappa was performed. In brief, 20 μl of soluble biotinylated ACE2 (from Sino Biological) at 1μg/mL, 10 μl of diluted serum (1/200 final dilution) and 20 μl of the 13 plex antigen-coupled beads premix are mixed. Following a 30-minute incubation on a shaker in the dark at room temperature, three wash steps were performed on a plate magnet using PBT. Subsequently, 40 μl of R-Phycoerythrin conjugated Streptavidin (from Jackson Immunoresearch) at 4 μg/mL was added into each well, and the plate was incubated on a shaker in the dark for 15 min, at room temperature. Lastly, three PBT washes were performed on a magnet and resuspended with 100 μl of PBT. Plates were read using a Luminex® MAGPIX® system and the median fluorescence intensity (MFI) was retained for analysis. To obtain a kinetic curve of the RBD-ACE2 binding inhibition for an individual sample, 7 points-2-fold serum dilution from 1/10 to 1/640 and one reference control point (without serum) were prepared in advance. The ACE2-RBD binding inhibition has been measured indirectly, as the MFI corresponds to fluorescence emitted by Streptavidin-phycoerythrin bound to biotinylated ACE2 antigen complex. Therefore, the MFI reading is inversely proportional to the concentration of neutralizing antibodies present in the serum.

### S-Fuse neutralization assay

U2OS-ACE2 GFP1–10 or GFP 11 cells, also termed S-Fuse cells, become GFP+ cells when they are productively infected with SARS-CoV-2. Cells tested negative for mycoplasma. Cells were mixed (at a 1:1 ratio) and plated at 8 × 10^3^ cells per well in a μClear 96-well plate (Greiner Bio-One). The indicated SARS-CoV-2 strains were incubated with sera at the indicated concentrations or dilutions for 15 min at room temperature and added to S-Fuse cells. The sera were heat inactivated for 30 min at 56 °C before use. Then, 18 hours later, cells were fixed with 2% paraformaldehyde, washed and stained with Hoechst (1:1,000 dilution; Invitrogen). Images were acquired with an Opera Phenix high-content confocal microscope (PerkinElmer). The GFP area and the number of nuclei were quantified using the Harmony software (PerkinElmer). The percentage of neutralization was calculated using the number of syncytia as the value with the following formula:

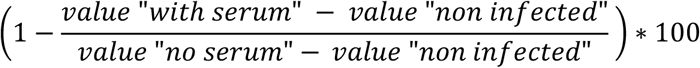

The neutralizing activity of each serum was expressed as the ED50 (reported in dilution units), which was calculated using a reconstructed curve based on the percentage of neutralization at different concentrations. An ED50 threshold of 30 has been defined below which sera samples have no neutralizing activity^18^.

### Data analysis

The ACE2-RBD binding inhibition was calculated for all 13 antigenic targets as a percentage of ACE2 maximum binding by dividing the MFI obtained for an individual serum by the MFI of the well without serum (only with ACE2, reference MFI), with the following calculation:

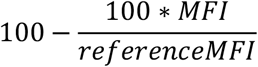

A threshold for positivity was established based on the analysis obtained with negative controls. We defined a threshold of 20% ACE2-RBD binding inhibition below which samples do not have neutralizing antibodies and are considered negative.

Microsoft Excel 2016 and R version 4.0.5. RStudio was used to run the analysis, with dplyr, readxl, tidy, ggplot2, openxlsx and stringr packages.

### Ethical approval

Collection of samples from the Orleans cohort had been approved by the Comité de Protection des Personnes Ile de France IV (NCT04750720). The study of the COVID-Oise cohort was registered with ClinicalTrials.gov (NCT04644159) and received ethical approval by the Comité de Protection des Personnes Nord Ouest IV. Several COVID-Oise participants participated in the CORSER studies in spring 2020, registered with ClinicalTrials.gov (NCT04325646) and approved by the Comité de Protection des Personnes Ile de France III. For all studies, participants did not receive any compensation. Informed consent was obtained from all participants, and parents provided informed consent for any children under the age of 18 years. For the nursing home residents who did not have full capacity to sign legal documents, informed consent was obtained from their relatives.

## Results

### Diversity of ACE2-RBD binding inhibition profiles after infection and/or vaccination

To validate the assay, we measured the dose response curves from five samples with differing immunological profiles according to infection and vaccination status. Five different characterized serum samples were tested, including a pre-pandemic negative pool, a positive pool of individuals infected at the beginning of the pandemic, two samples from individuals vaccinated with one or two doses respectively, as well as a sample from an infected and vaccinated (1 dose) individual.

At the lowest serum dilution the percentage of inhibition of the ACE2-RBD binding is zero for the negative pre-pandemic pool (Fig. 1). The ACE2-RBD binding inhibition increase over 45% for RBD WT after one single dose of vaccine. A higher ACE2-RBD binding inhibition is observed for the double vaccinated individual sample compared to the positive pool, with an inhibition percentage of 90% and 70% respectively. We note that for these last four samples the percentage of inhibition of ACE2-RBD binding decreases for most VOCs, especially for Omicron. For the vaccinated plus infected individual (hybrid immunity), we observed an ACE2-RBD binding inhibition around 100%, not only for the ancestral form but also for the VOCs (except for Beta and Omicron variant for which the maximum value is below 90% inhibition). The large inter-individual variation observed for these five representative samples, due to their different serological status, enabled us to validate the test.

**Figure 1:**
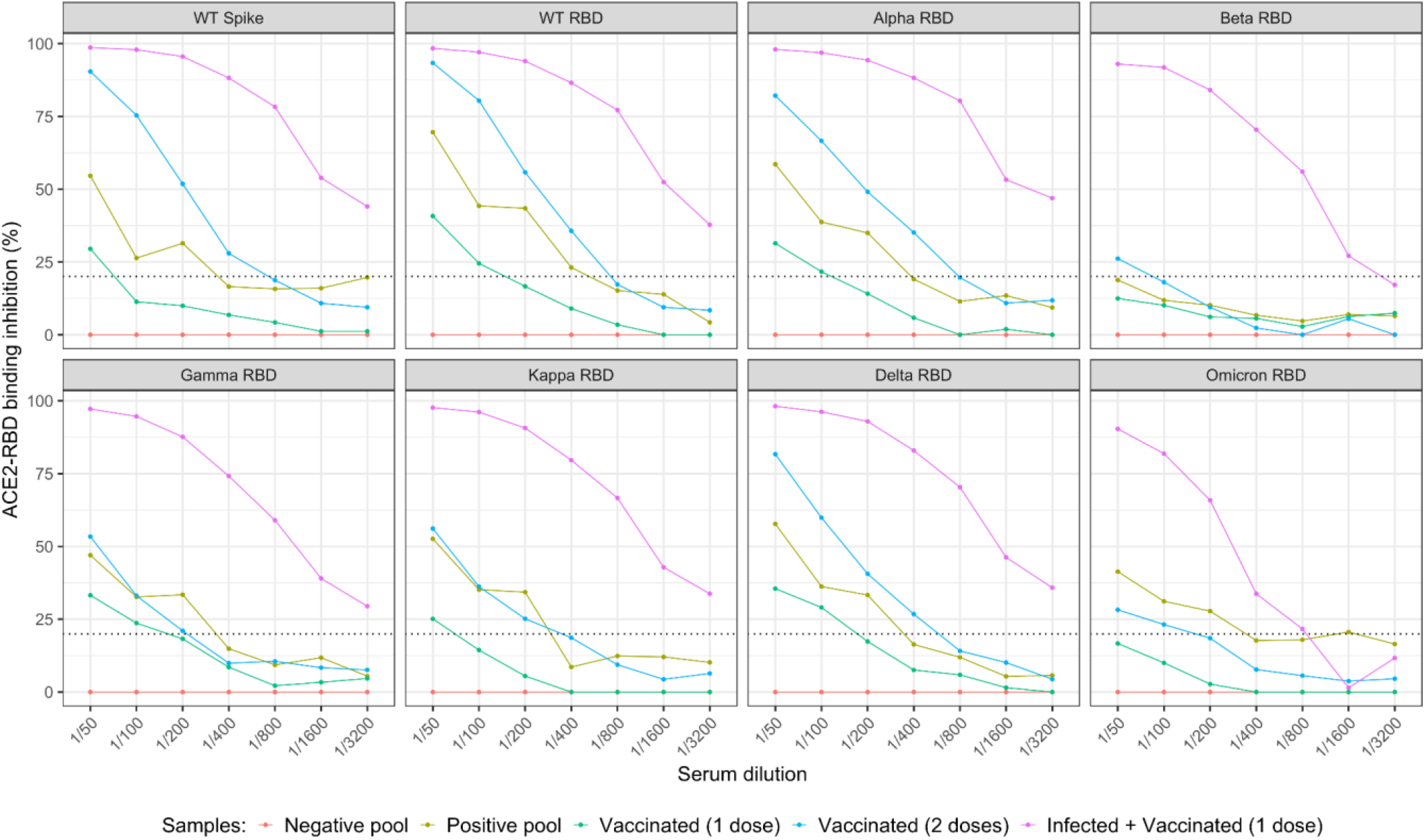
Inhibition profiles of SARS-CoV-2 RBD-ACE2 binding for five representative samples of differing status (uninfected, infected and/or vaccinated). A bead-based multiplex assay using three characterized vaccinated serum samples: 1 dose (green), 2 doses (blue) and 1 dose plus SARS-CoV-2 infection (pink) was used. A positive pool of plasma from infected individuals (yellow) and a negative pre-pandemic pool (red) was used. Results are shown for WT Spike and RBD antigens, as well as RBD antigens of SARS-CoV-2 variants Alpha, Beta, Gamma, Kappa, Delta and Omicron (lineage BA.1). A 7-point 2-fold titration was performed for all samples. The plots show the percentage of RBD-ACE2 inhibition according to serum dilution. The horizontal dotted line represents the threshold of the assay.

### Association between ACE2-RBD binding inhibition, live virus neutralization and IgG antibody levels

We investigated the association between the ACE2-RBD binding inhibition data from our surrogate virus neutralization test and the ED50 data from the live virus neutralization assay. We have defined a threshold of 20% ACE2-RBD binding inhibition for our assay and an ED50 threshold of 30. Values below these thresholds indicate that antibodies present in the serum are unable to inhibit the binding, therefore the serum has limited neutralizing effect. We observe strong association between the ED50 value and ACE2-RBD binding inhibition, especially for RBD WT, Alpha, Beta and Delta variants (R^2^ between 0.68 and 0.73). When we restrict to samples with measured neutralization activity (ED50 > 30), we observe a linear association and noticeably stronger correlation between 0.66, and 0.79 (Fig. 2 panel A). The strong correlation between both assays allows us to validate our surrogate virus neutralization assay, which aims to estimate the capacity of antibodies present in the serum to inhibit the ACE2-RBD binding and thus prevent virus entry.

**Figure 2:**
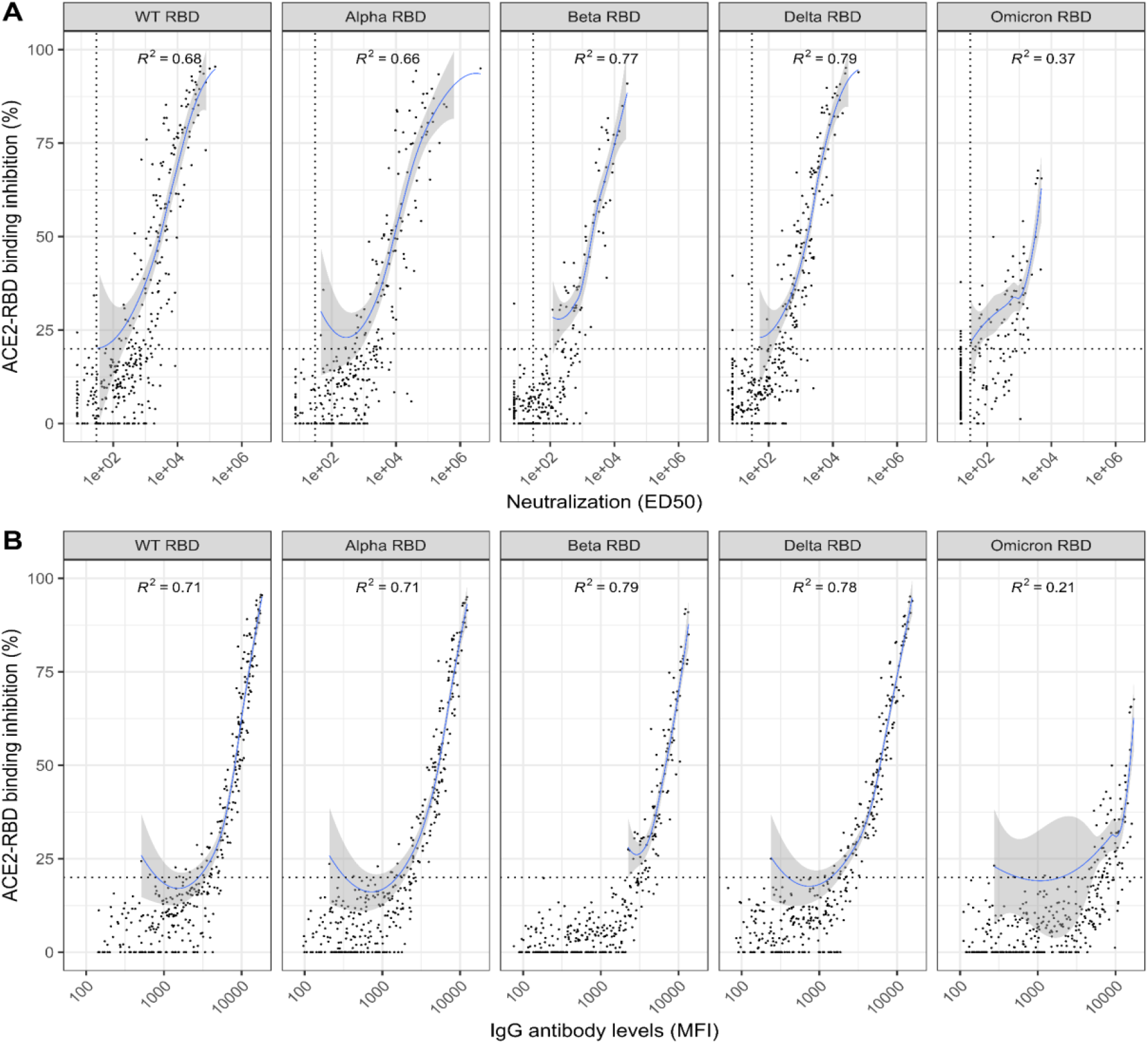
A) Association between ACE2-RBD binding inhibition and EC50 live virus neutralization. B) Association between ACE2-RBD binding inhibition and IgG antibody levels. Samples from vaccinated and/or infected individuals were used (n=412). Results are shown for WT RBD and VOC RBDs. The horizontal dotted lines on panel A and B correspond to our in-house assay threshold. The vertical dotted lines on panel A correspond to the live virus neutralization assay threshold.

Subsequently, we study if there was a correlation between IgG antibody levels (MFI) and the ACE2-RBD binding inhibition. For this purpose, we used a multiplex serology test to measure IgG levels and compared to the ACE2-RBD binding inhibition data. The correlation between IgG antibody levels and their neutralizing capacities is non-linear (Fig. 2, panel B). For ACE2-RBD binding inhibition >20% we observe an approximately linear relationship, with correlation coefficients in the range 0.71 – 0.79, with the exception of Omicron which had a lower correlation of 0.21 owing to lower levels of binding inhibition.

### Neutralizing antibody levels in individuals followed one-year post-vaccination

A cohort of vaccinated individuals was followed with samples collected from week 2 after the first dose and continued until 6 months after the second dose.

In samples taken from individuals just after administration of the first vaccine dose, negligible binding inhibition and ED50 neutralization value (Fig. 3) was observed. The ED50 value and the binding inhibition increases considerably for all variants after second dose, especially WT RBD, Delta RBD and Alpha RBD, with a binding inhibition of 49.1%, 41% and 36.1% respectively. It is worth noting that for the live virus neutralization results, a higher titer of neutralizing antibodies is directed towards Alpha RBD and subsequently to WT RBD and Delta RBD. This confirms that the neutralizing antibodies produced after vaccination cross-react with several variants of the virus.

**Figure 3:**
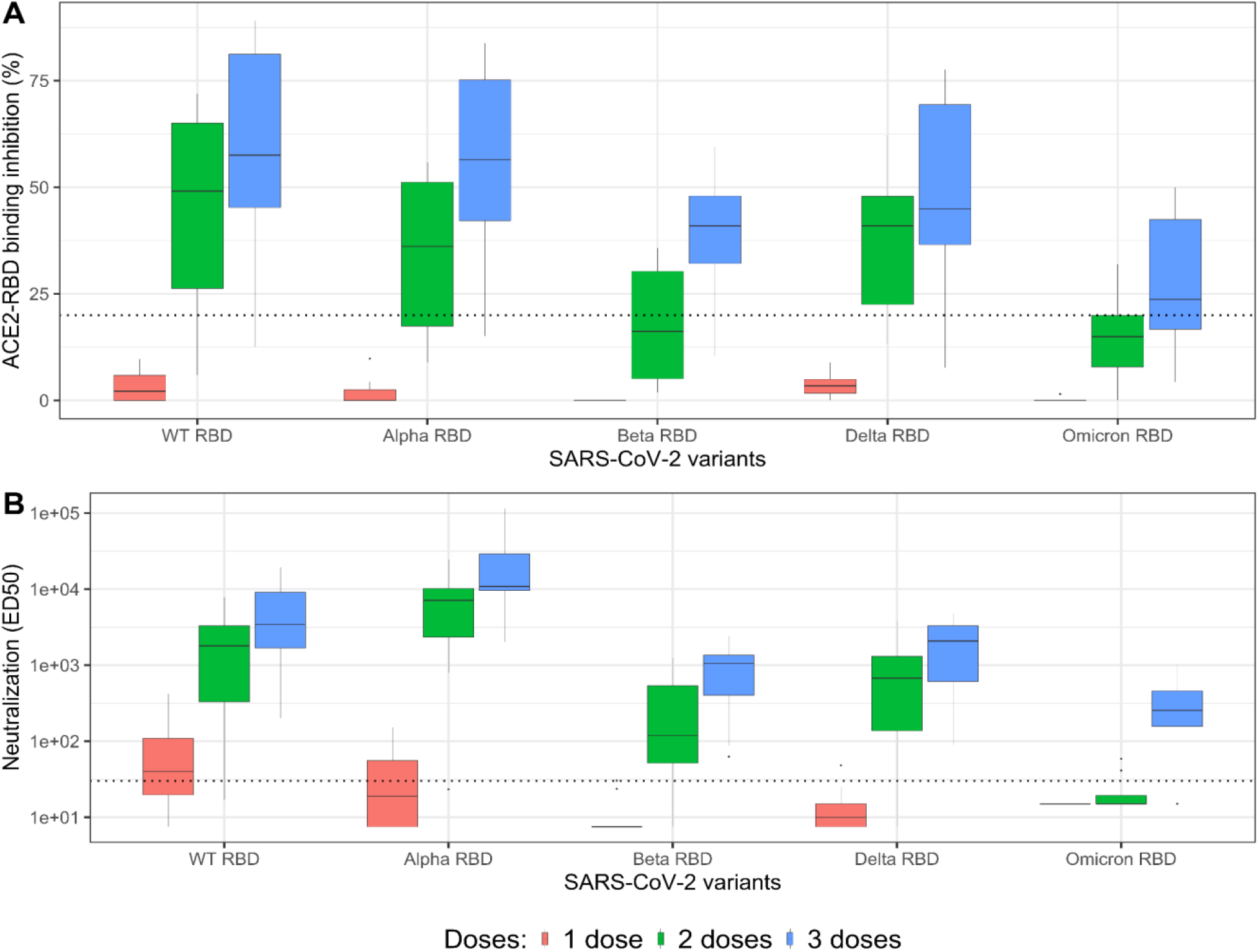
ACE2-RBD binding inhibition and ED50 neutralization according to the number of Pfizer/BioNTech BNT162b2 vaccine doses. Samples from 11 individuals were collected three weeks after each dose of vaccine (n=36). Results from WT RBD and VOC RBDs are shown. On top are shown the results obtained with the multiplex surrogate virus neutralization test and on the bottom live virus neutralization assay. The dotted lines correspond to the respective thresholds.

As described previously, COVID-19 vaccines are not equally effective against all variants^16^. Following a third dose, a higher antibody neutralizing capacity is noticeable for RBD WT, Alpha, and Delta with a median ACE2-RBD binding inhibition of 57.5%, 56.5% and 44.9% respectively. Followed by a significant decrease for the latest VOC: Omicron, with a median of 23.7%, consistent with lower vaccine efficacy. Similar results are observed for the live virus neutralization assay, an increase in the ED50 value is observed after the booster dose, suggesting that vaccination induces the production of neutralizing antibodies against several variants, especially against RBD WT and Alpha. On the contrary, for the vaccination regimens considered, the median neutralization activity for the SARS-CoV-2 Omicron variant is under the thresholds of ED50 = 30, suggesting that there are limited levels of anti-RBD Omicron neutralizing antibodies present in the serum.

The kinetics of neutralizing antibodies and ACE2-RBD binding inhibition followed similar patterns over time. A peak of neutralizing antibody titers and ACE2-RBD binding inhibition appears around week 4 and week 36 post vaccination, corresponding to 4 weeks after the first dose and booster dose respectively (Fig. 4). This representation showed a rapid decline of the ACE2-RBD binding inhibition over 20 weeks. Similarly, a decrease in the ED50 value after week 6 is observed for the live virus neutralization assay, however this decrease in neutralizing antibodies is not as pronounced. Moreover, anti-RBD neutralizing antibodies decreases considerably four and a half months after second vaccine dose, suggesting that COVID-19 vaccination will probably have short-lasting protective effect.

**Figure 4:**
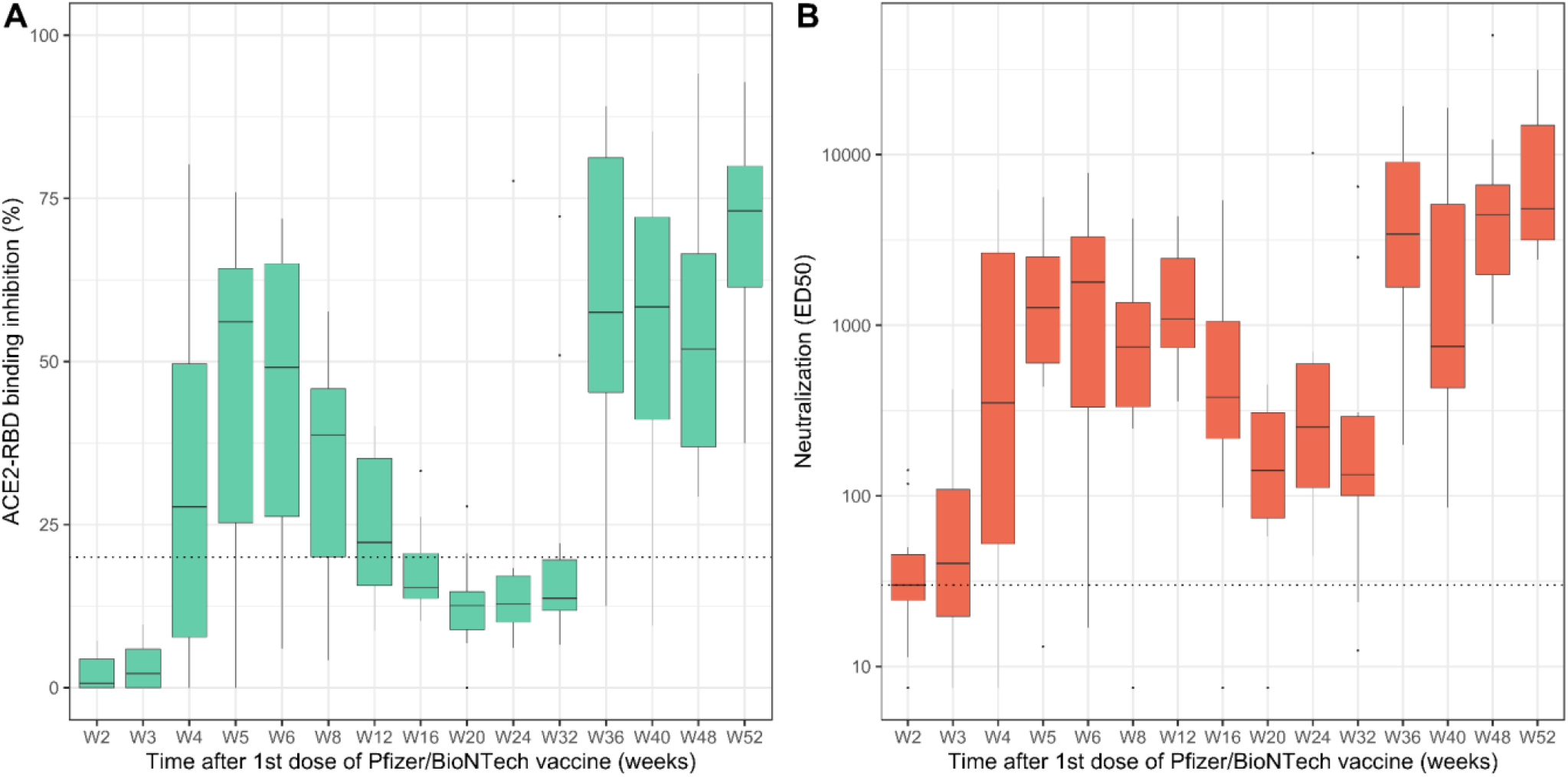
Kinetics of ACE2-RBD binding inhibition and neutralization titer overtime following Pfizer/BioNTech BNT162b2 vaccination. Samples from 11 individuals were collected frequently after their first dose. On the left the results obtained with the surrogate virus neutralization assay and on the right live virus neutralization assay. Results are shown for RBD WT antigen. The dotted lines correspond to the respective thresholds.

### Protection estimates

The SARS-CoV-2 surrogate live virus neutralization assay enables us to assess protection estimates against breakthrough infection in large population studies. This assay was applied to samples from a longitudinal French cohort study from the town of Crépy-en-Valois (Covid-Oise study), where sera samples were collected from approximately 515 individuals in December 2021 (Session 3) and April 2022 (Session 4). During this time, the Omicron (lineages BA.1 and BA.2) wave swept through France, infecting large numbers of individuals. Most individuals had received between 1 and 2 doses of vaccine.

By measuring anti-N IgG responses, we identified individuals who were infected before Session 3 or Session 4. Moreover, our analysis enabled the identification of individuals who experienced reinfection between the two sampling periods, by detecting individuals who exhibited a four-fold increase in anti-N antibody levels (Table 1). This analysis allows the categorization of individuals into five subgroups based on their serostatus, each denoted by a short name in Table 1 and Fig. 5. Individuals who tested negative at Session 3 and Session 4 are classified as “neg_neg”. Those who were negative in Session 3 but positive in Session 4 are termed “neg_pos”. Individuals who tested positive at Session 3 but subsequently sero-reverted to negative by Session 4 are classified as “pos_neg”. Those who tested positive at both sessions without evidence of new infection between the two cross-sections are labeled as “pos_pos”. Lastly, individuals who tested positive at Session 3 and were identified as being reinfected between the two sessions are designated as “pos_boost”. In this subgroup, the significant rise of the median anti-N antibodies levels from 28 to 840 (expressed in x10-5) between the two sessions strongly indicates a re-infection event.

**Table 1:** Overview of the five different serostatus subgroups within the sample set, including the number and percentage of individuals in each subgroup. The median and interquartile range of anti-N antibody levels for sessions 3 and 4, are expressed in RAU. The median and interquartile range of ACE2-RBD binding inhibition for Wuhan and Omicron strains measured at Session 3 are detailed. The median age, number of females, and vaccination status (number of vaccine doses) are also provided for each subgroup.

| categories |  | neg_neg | pos_neg | pos_pos | neg_pos | pos_boost |
| --- | --- | --- | --- | --- | --- | --- |
| count n (%) |  | 94 (18.3 %) | 22 (4.3 %) | 198 (38.4 %) | 107 (20.8 %) | 94 (18.3 %) |
| anti-NP IgG median *<br>[IQR]<br>*expressed in $\times 10^{-5}$ | Session 3 | 5.8 [4.6;7.4] | 13[12;18] | 65 [32;150] | 5.7 [4.2;7.6] | 28 [15;64] |
|  | Session 4 | 5.3 [4.2;6.9] | 7.2 [5.4;8.4] | 37 [20;72] | 82 [32; 210] | 840 [250;2500] |
| ACE2-RBD binding<br>inhibition median at<br>Session 3 [IQR] | Wuhan strain | 26.8%<br>[12.7;47.4] | 45.7%<br>[32.6;65.4] | 71.4%<br>[31.5;90.4] | 24.5%<br>[12.9;41.3] | 57.2%<br>[22.3;83.3] |
|  | Omicron strain | 14.3%<br>[8.5;18.2] | 19% [14.2;21.9] | 24.1%<br>[13.7;41.2] | 14.7% [9.8;20] | 17.8%<br>[11.9;30.1] |
| age [min;max] |  | 50 [9;93] | 52 [17;80] | 52 [9;98] | 39 [6;85] | 41 [7;94] |
| female count n (%) |  | 58 (61.7%) | 17 (77.3%) | 128 (64.6%) | 64 (59.8%) | 70 (74.5%) |
| Vaccination status<br>count n (%) | unvaccinated | 7 (7.4%) | 1 (4.5%) | 22 (11.1%) | 19 (17.8%) | 17 (18.1%) |
|  | one dose | 8 (8.5%) | 9 (40.9%) | 95 (48%) | 10 (9.3%) | 43 (45.7%) |
|  | two doses | 71 (75.5%) | 12 (54.5%) | 58 (29.3%) | 72 (67.3%) | 29 (30.9%) |
|  | three doses | 7 (7.4%) | 0 (0%) | 23 (11.6%) | 5 (4.7%) | 5 (5.3%) |
|  | unknown | 1 (1.1%) | 0 (0%) | 0 (0%) | 1 (0.9%) | 0 (0%) |

**Figure 5:**
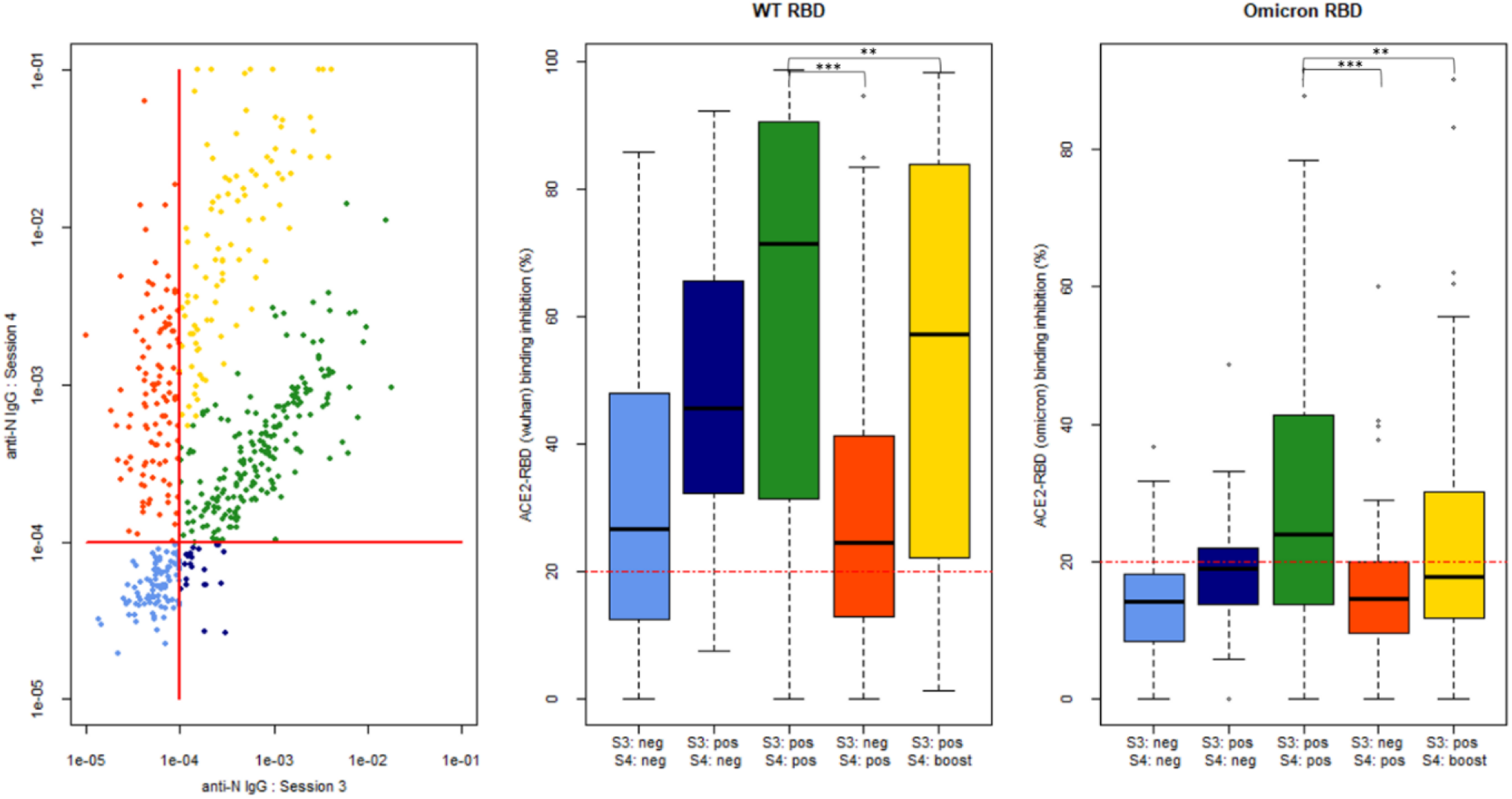
Panel A) Levels of anti-N IgG from Covid-Oise cohort Session 3 (December 2021) and Session 4 (April 2022) are illustrated for each individual, ensuring matched comparisons between sessions. Each category is consistently color-coded across the different panels in accordance with the descriptions provided previously. In light blue are individuals, who were negative in Session 3 and Session 4 (“neg_neg”). In red are individuals negative in Session 3 and positive in Session 4 (“neg_pos”). In green are individuals who were positive at both sessions, but without evidence of new infection between the two cross-sections (“pos_pos”). In yellow are individuals positive at Session 3 and identified as being reinfected between the two cross-sections. In dark blue are individuals who were positive at Session 3 but then sero-reverted to negative at Session 4. The horizontal and vertical red lines represent the threshold for seropositivity. Panel B) The ACE2-RBD binding inhibition for the WT variant at Session 3 among the previously presented subgroups. The red horizontal dotted line represents the threshold of our assay. Panel C) The ACE2-RBD binding inhibition for the Omicron variant at Session 3 among the previously described subgroups. The red horizontal dotted line represents the threshold of our assay.

We investigated correlates of protection by analyzing ACE2-RBD WT and Omicron binding inhibition measurements obtained at Session 3 along with the serostatus defined at both Sessions (Fig. 5). The first key comparison was between groups with no evidence of infection before Session 3 (anti-N IgG sero-negative). Both the “neg_neg” and “neg_pos” groups had high levels of ACE2-RBD WT binding inhibition, consistent with high levels of vaccine-induced immunity. There was no significant difference in ACE2-RBD WT binding inhibition between these groups (P value = XX; wilcoxon test), consistent with the hypothesis that ACE2-RBD WT binding inhibition is not associated with protection from infection with the SARS-CoV-2 Omicron Variant in vaccinated individuals. Furthermore, all individuals who were uninfected before Session 3 had very low levels of ACE2-RBD Omicron binding inhibition, so it was not possible to detect significant associations between Omicron specific humoral immune responses and protection from infection.

The second key comparison is between groups with evidence of infection before Session 3 (anti-N IgG sero-positive). Both the “pos_pos” and “pos_boost” groups had high levels of ACE2-RBD WT binding inhibition. The “pos_pos” group had significantly higher ACE2-RBD WT binding inhibition that the “pos_boost” group (P value = 0.008), consistent with the hypothesis that in individuals with hybrid immunity, greater ACE2-RBD WT binding inhibition is associated with protection from infection. Although Omicron specific ACE2-RBD binding inhibition levels were substantially lower, we still observed a significant association between ACE2-RBD Omicron binding inhibition and protection from infection (P value = 0.001). These results indicates that RBD Omicron binding inhibition is not associated with protection against Omicron infection in individuals who have acquired immunity solely through vaccination. However, for individuals with hybrid immunity (due to both vaccination and previous infection), higher RBD Omicron binding inhibition is associated with increased protection against an Omicron infection.

## Discussion

To investigate the neutralizing capacity of antibodies against SARS-CoV-2 variants, we developed a multiplex surrogate virus neutralization test based on ACE2-RBD binding inhibition. This allowed us to develop an accurate and rapid high-throughput multiplex test that could adapt easily as numerous SARS-CoV-2 variants emerged. Additionally, our test overcomes the main disadvantages of conventional virus neutralization assay, since no virus, no live cells, no large volume of sera and limited biosafety requirements are needed. The analysis of well-characterized sera samples from infected and/or vaccinated subjects showed a strong correlation between the surrogate virus neutralization test and the gold standard live virus neutralization assay.

We studied vaccinated individuals to gain deeper insights into antibody kinetics following vaccination. Prior research has suggested a characteristic pattern in antibody levels: a peak three weeks after infection or vaccination is observed, followed by a rapid decline in subsequent months, and then a slower decrease eight months post immunization^19–21^. Our findings align with this pattern, revealing that both binding and neutralizing antibodies follows a similar kinetic post-vaccination. This suggests that the neutralizing capacity of antibodies is closely correlated with antibody levels and their maturation. Notably, a third vaccine dose was associated with significantly higher ACE2-RBD binding inhibition compared to two doses, indicating that a booster dose enhances protection against all VOCs. This suggests that COVID-19 vaccination may confer short-lasting protective effects, necessitating additional booster doses for certain population groups as elderly or immunocompromised individuals^22,23^.

Additionally, our study provides valuable insights into the neutralizing antibody response to various SARS-CoV-2 variants and its role in providing protection against Omicron breakthrough infections within the vaccinated population. We noted among vaccinated individuals, that the ACE2-RBD binding inhibition against RBD variants, especially Omicron, is reduced compared to the ancestral virus. As described in the literature, the accumulation of mutations within the Omicron variant has an impact on the binding affinity to ACE2, which gives a greater chance to evade host immunity, enhancing transmissibility^24^. Studies have shown that mutations on Omicron RBD result in stronger binding to ACE2, therefore antibodies elicited by vaccine-acquire immunity showed reduced ACE2-RBD binding inhibition and neutralization potential^25^. Indeed, RBD is responsible for the attachment to ACE2 human cell receptor, thus high avidity and neutralizing anti-RBD antibodies plays an essential part in the prevention of infection by blocking this interaction.

In our study, samples from a longitudinal population cohort collected in December 2021 and April 2022, were used to measure immunity to breakthrough SARS-CoV-2 infection. Results shown that the ACE2-RBD binding inhibition from individuals infected prior to Session 3 and re-infected between both Sessions (probably with the Omicron variant), showed lower levels of neutralizing antibodies compared to individuals infected prior to Session 3 and without any sign of a new infection. These individuals who experienced reinfection were likely more susceptible due to the fact that their levels of neutralizing antibodies were lower at Session 3 (52.7% and 17.8% for Wuhan and Omicron variant respectively). Suggesting that two doses of the vaccine may not provide adequate protection against the Omicron variant for some individuals, underscoring the importance of implementing booster doses. This is particularly crucial as we observed breakthrough infections occurring despite initial infection. However, our study also indicates that individuals with high levels of antibodies from hybrid immunization, such as those induced by previous infection and subsequent vaccination, demonstrate enhanced protection against emerging variants like Omicron. This underscores the importance of continually assessing the evolution of SARS-CoV-2 and its variants, along with evaluating the effectiveness of vaccine induced anti-RBD antibodies against diverse strains. Such efforts are essential for better understanding the protective immune response.

While our assay provided valuable insights, it is crucial to acknowledge its limitations, as it only allows us to measure the MFI of the Streptavidin phycoerythrin bound to the biotinylated ACE2 receptor. We do not directly read the MFI of antibodies bound to the antigens coupled beads. Consequently, the isotype of the neutralizing antibodies in the serum is unknown. Several studies demonstrated that specific mucosal IgA antibodies have a crucial role in early virus neutralization^26^. This is a notable limit of our study as we only optimized and performed this test on serum samples, since we intended to have a global view of neutralizing antibody levels and the protection given by vaccination. It could be contemplated in the future to conduct our assay using nasopharyngeal or saliva samples, in the cases of infection acquired immunity. It is important to note that these experiments have been done with a limited sample diversity, as the major global vaccination campaign, made it difficult to do serological follow-ups on unvaccinated individuals infected by new SARS-CoV-2 variants.

## Conclusion

The development of this multiplex surrogate virus neutralization test based on ACE2-RBD binding provides accurate information on neutralizing antibodies levels that can allow us to better understand the kinetics, their functionality and the protection provided at population level. Therefore, we can apply this high-throughput tool to SARS-CoV-2 emerging variants to estimate functional protection conferred by vaccination or infection. This assay enables us to assess clinical protection against SARS-CoV-2 variants and evaluate the effectiveness of vaccines or antibody therapeutics, providing essential information to monitor the pandemic’s evolution.

## Author contributions

EB developed the assay, analysed samples, and wrote the first draft of the manuscript. EB and MW analysed the data. LG, FD, SP and JR supported assay development. DP and TB preformed the neutralization assay. AF, LP and OS provided access to samples. MW designed the study.

## Acknowledgements

The authors thank the participants who agreed to participate into the different studies and the medical and paramedical teams who were involved in sample and data collection. They thank the teams of Crépy-en-Valois town hall and the director and technical services of the local hospital for their help in implementing the COVID-Oise study. They also thank the Institut Pasteur’s Investigation Clinique et Accès aux Ressources Biologiques team for sample management and Institut Pasteur’s Pôle de Coordination de la Recherche Clinique for support with regulatory processes.

## Data availability

All data produced in the present study are available on request to the authors.

## References

1. Gorkhali, R. et al. Structure and Function of Major SARS-CoV-2 and SARS-CoV Proteins. Bioinforma. Biol. Insights 15, 11779322211025876 (2021).

2. V’kovski, P., Kratzel, A., Steiner, S., Stalder, H. & Thiel, V. Coronavirus biology and replication: implications for SARS-CoV-2. Nat. Rev. Microbiol. 19, 155–170 (2021).

3. Scialo, F. et al. ACE2: The Major Cell Entry Receptor for SARS-CoV-2. Lung 198, 867–877 (2020).

4. Min, L. & Sun, Q. Antibodies and Vaccines Target RBD of SARS-CoV-2. Front. Mol. Biosci. 8, (2021).

5. Marot, S. et al. Rapid decline of neutralizing antibodies against SARS-CoV-2 among infected healthcare workers. Nat. Commun. 12, 844 (2021).

6. Burton, D. R., Williamson, R. A. & Parren, P. W. H. I. Antibody and Virus: Binding and Neutralization. Virology 270, 1–3 (2000).

7. Piccoli, L. et al. Mapping Neutralizing and Immunodominant Sites on the SARS-CoV-2 Spike Receptor-Binding Domain by Structure-Guided High-Resolution Serology. Cell 183, 1024-1042.e21 (2020).

8. Khoury, D. S. et al. Neutralizing antibody levels are highly predictive of immune protection from symptomatic SARS-CoV-2 infection. Nat. Med. 27, 1205–1211 (2021).

9. Earle, K. A. et al. Evidence for antibody as a protective correlate for COVID-19 vaccines. Vaccine 39, 4423–4428 (2021).

10. Cosar, B. et al. SARS-CoV-2 Mutations and their Viral Variants. Cytokine Growth Factor Rev. 63, 10–22 (2022).

11. Lopez, E. et al. Simultaneous evaluation of antibodies that inhibit SARS-CoV-2 variants via multiplex assay. JCI Insight 6, (2021).

12. Tan, C. W. et al. A SARS-CoV-2 surrogate virus neutralization test based on antibody-mediated blockage of ACE2-spike protein-protein interaction. Nat. Biotechnol. 38, 1073–1078 (2020).

13. Woudenberg, T. et al. Estimated protection against COVID-19 based on predicted neutralisation titres from multiple antibody measurements in a longitudinal cohort, France, April 2020 to November 2021. Eurosurveillance 28, 2200681 (2023).

14. Fontanet, A. et al. SARS-CoV-2 infection in schools in a northern French city: a retrospective serological cohort study in an area of high transmission, France, January to April 2020. Eurosurveillance 26, 2001695 (2021).

15. Planas, D. et al. Considerable escape of SARS-CoV-2 Omicron to antibody neutralization. Nature 602, 671–675 (2022).

16. Woudenberg, T. et al. Humoral Immunity to SARS-CoV-2 and Inferred Protection from Infection in a French Longitudinal Community Cohort. (2022).

17. Rosado, J. et al. Multiplex assays for the identification of serological signatures of SARS-CoV-2 infection: an antibody-based diagnostic and machine learning study. Lancet Microbe 2, e60–e69 (2021).

18. Bruel, T. et al. Serum neutralization of SARS-CoV-2 Omicron sublineages BA.1 and BA.2 in patients receiving monoclonal antibodies. Nat. Med. (2022) doi:10.1038/s41591-022-01792-5.

19. Bates, T. A. et al. Neutralization of SARS-CoV-2 variants by convalescent and BNT162b2 vaccinated serum. Nat. Commun. 12, 5135 (2021).

20. Reifer, J., Hayum, N., Heszkel, B., Klagsbald, I. & Streva, V. A. SARS-CoV-2 IgG antibody responses in New York City. Diagn. Microbiol. Infect. Dis. 98, 115128 (2020).

21. Pelleau, S. et al. Kinetics of the Severe Acute Respiratory Syndrome Coronavirus 2 Antibody Response and Serological Estimation of Time Since Infection. J. Infect. Dis. 224, 1489–1499 (2021).

22. Levin Einav G. et al. Waning Immune Humoral Response to BNT162b2 Covid-19 Vaccine over 6 Months. N. Engl. J. Med. 385, e84 (2021).

23. Samanovic, M. I. et al. Vaccine-Acquired SARS-CoV-2 Immunity versus Infection-Acquired Immunity: A Comparison of Three COVID-19 Vaccines. Vaccines 10, 2152 (2022).

24. Chen, L.-L. et al. Impact of Severe Acute Respiratory Syndrome Coronavirus 2 (SARS-CoV-2) Variant-Associated Receptor Binding Domain (RBD) Mutations on the Susceptibility to Serum Antibodies Elicited by Coronavirus Disease 2019 (COVID-19) Infection or Vaccination. Clin. Infect. Dis. 74, 1623– 1630 (2022).

25. Lupala, C. S., Ye, Y., Chen, H., Su, X.-D. & Liu, H. Mutations on RBD of SARS-CoV-2 Omicron variant result in stronger binding to human ACE2 receptor. Biochem. Biophys. Res. Commun. 590, 34–41 (2022).

26. Sterlin, D. et al. IgA dominates the early neutralizing antibody response to SARS-CoV-2. Sci. Transl. Med. 13, eabd2223 (2021).

